# Genetics of posttraumatic stress disorder and cardiovascular conditions using Life’s Essential 8, Electronic Health Records, and Heart Imaging

**DOI:** 10.1101/2024.08.20.24312181

**Authors:** Jie Shen, Wander Valentim, Eleni Friligkou, Cassie Overstreet, Karmel Choi, Dora Koller, Christopher J. O’Donnell, Murray B. Stein, Joel Gelernter, Posttraumatic Stress Disorder Working Group of the Psychiatric Genomics Consortium, Haitao Lv, Ling Sun, Guido J. Falcone, Renato Polimanti, Gita A. Pathak

## Abstract

**BACKGROUND:** Patients with post-traumatic stress disorder (PTSD) experience higher risk of adverse cardiovascular (CV) outcomes. This study explores shared loci, and genes between PTSD and CV conditions from three major domains: CV diagnoses from electronic health records (CV-EHR), cardiac and aortic imaging, and CV health behaviors defined in Life’s Essential 8 (LE8).

**METHODS:** We used genome-wide association study (GWAS) of PTSD (N=1,222,882), 246 CV diagnoses based on EHR data from Million Veteran Program (MVP; N=458,061), UK Biobank (UKBB; N=420,531), 82 cardiac and aortic imaging traits (N=26,893), and GWAS of traits defined in the LE8 (N = 282,271 ∼ 1,320,016). Shared loci between PTSD and CV conditions were identified using local genetic correlations (rg), and colocalization (shared causal variants). Overlapping genes between PTSD and CV conditions were identified from genetically regulated proteome expression in brain and blood tissues, and subsequently tested to identify functional pathways and gene-drug targets. Epidemiological replication of EHR-CV diagnoses was performed in AllofUS cohort (AoU; N=249,906).

**RESULTS:** Among the 76 PTSD-susceptibility risk loci, 33 loci exhibited local rg with 45 CV-EHR traits (|rg|≥0.4), four loci with eight heart imaging traits(|rg|≥0.5), and 44 loci with LE8 factors (|rg|≥0.36) in MVP. Among significantly correlated loci, we found shared causal variants (colocalization probability > 80%) between PTSD and 17 CV-EHR (in MVP) at 11 loci in MVP, that also replicated in UKBB and/or other cohorts. Of the 17 traits, the observational analysis in the AoU showed PTSD was associated with 13 CV-EHR traits after accounting for socioeconomic factors and depression diagnosis. PTSD colocalized with eight heart imaging traits on 2 loci and with LE8 factors on 31 loci. Leveraging blood and brain proteome expression, we found 33 and 122 genes, respectively, shared between PTSD and CVD. Blood proteome genes were related to neuronal and immune processes, while the brain proteome genes converged on metabolic and calcium-modulating pathways (FDR p <0.05). Drug repurposing analysis highlighted *DRD2, NOS1, GFAP, and POR* as common targets of psychiatric and CV drugs.

**CONCLUSION:** PTSD-CV comorbidities exhibit shared risk loci, and genes involved in tissue-specific regulatory mechanisms.

## 1 Introduction

Given that cardiovascular (CV)-related outcomes, including diseases and risk factors are the leading cause of morbidity and mortality worldwide ^1^, it is imperative to extend our understanding of associations beyond traditionally known CV risk factors. Recently, the American Heart Association recognized the influence of psychological stress on adverse CV health ^2^. Posttraumatic stress disorder (PTSD) is considered a stress-related mental disorder with a lifetime prevalence ranging from 2% to 25% ^3^. PTSD and CV diseases are highly comorbid. Specifically, PTSD has been associated with CVD ^4^, hypertension ^5^, diabetes ^6^, ischemic heart disease ^7^, stroke ^8^, carotid intima-media thickness ^9^, and coronary artery disease ^10^. PTSD and CVD have been reported to share several pathophysiological features. For instance, patients with PTSD have higher adrenergic activity both at baseline and after being subjected to stress-inducing situations ^11^, imposing a chronically increased burden on both the heart and the circulatory system. High adrenergic states are also associated with CVD symptoms, including increased heart rate and acutely increased blood pressure ^12^. Additionally, PTSD can increase the risk of CVDs by increasing the risk of dyslipidemia and diabetes, although the biology of this association is not yet fully understood ^13, 14^.

Both PTSD and CVD have substantial genetic components, with heritability estimates of 30-40% for PTSD ^13, 15^ and 15-57% for hypertension, 26% for heart failure (HF), and 40-60% for coronary artery disease ^16–20^. Recently, PTSD polygenic risk has been associated with several cardiovascular symptoms and disorders ^21^ and showed potential genetic causality towards cardiac arrhythmias ^21^, ischemic stroke ^22^, coronary artery disease, and hypertension ^23^. However, several critical gaps remain in understanding the shared genetics between PTSD and CVD which were highlighted by the experts from the American Heart Association (AHA) and the National Heart, Lung, and Blood Institute ^24^ . In this study, we address some of these gaps by leveraging GWAS (genome-wide association study) data to study genetic overlap between PTSD ^25^ and CV outcomes from three different domains including EHR diagnoses, heart imaging ^26^ and Life’s Essential 8 (LE8) key measures for improving and maintaining – evidence-based cardiovascular health as factors and behaviors recently outlined by the American Heart Association ^2^ . Specifically, we aimed to (i) identify loci that are shared between PTSD and CV conditions, (ii) test the specificity of PTSD-CVD comorbidity accounting for socioeconomic factors (BMI, smoking, deprivation index) and diagnosis of depression, and (iii) infer biological mechanisms underlying PTSD and CV outcomes based on tissue-specific transcriptomic regulation and proteomic gene-associations that overlap between PTSD and CVD traits (Figure 1).

**Figure 1:**
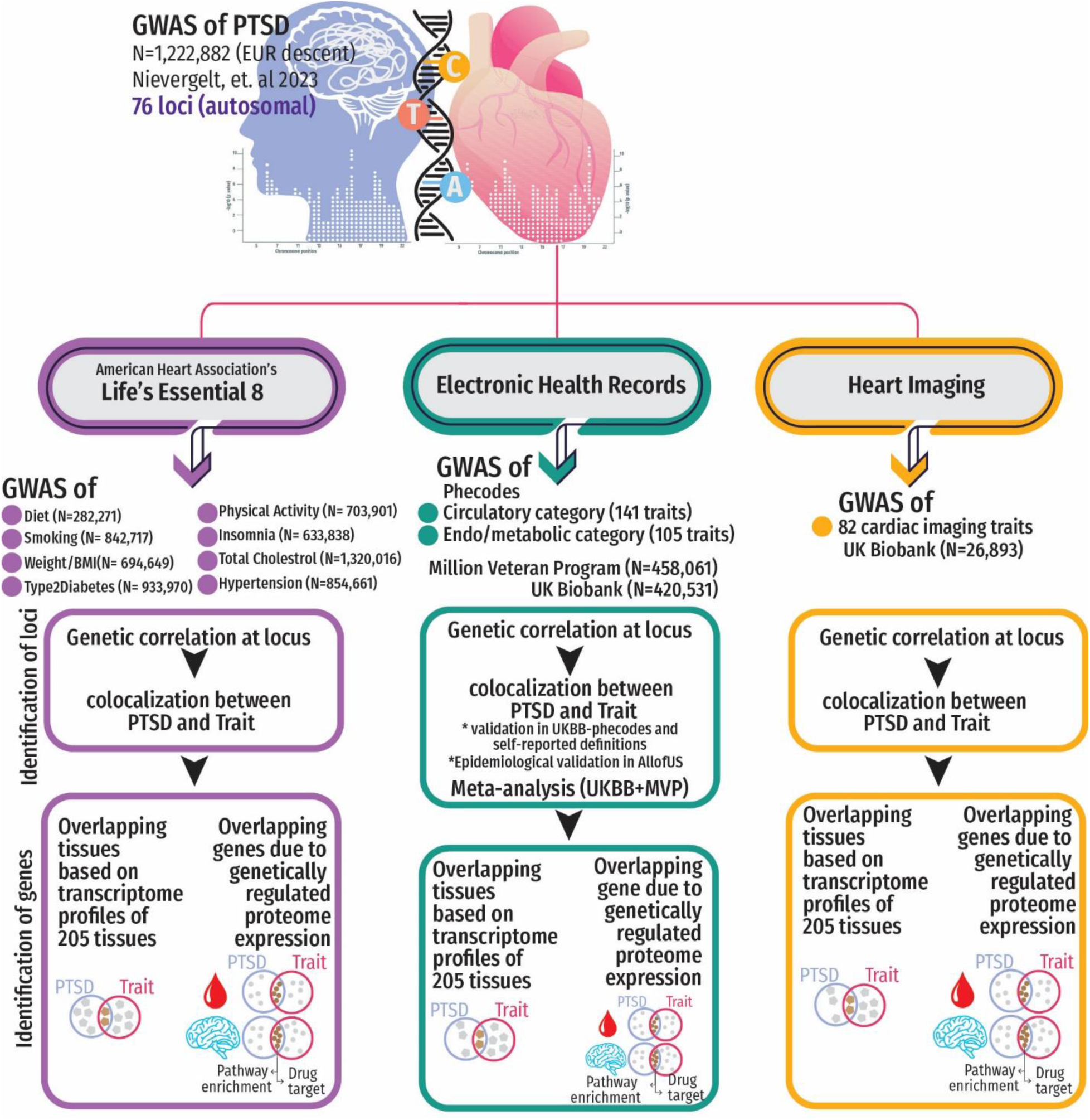
Study Design: We investigated loci shared between posttraumatic stress disorder (PTSD) and cardiovascular (CV)-related traits, including American Heart Association’s Life Essential 8 factors, CV diagnoses derived from electronic health records (EHR), and cardiac imaging phenotypes. After identifying genetically correlated loci between PTSD and CV conditions, we investigated shared causal variants. For the EHR-based CV diagnoses, we performed replication of shared causal variants in UK Biobank and a follow-up analysis in All of US Research Program. The traits with evidence of PTSD-CV shared causal variants were tested with respect to tissue-specific transcriptomic and proteomic profiles. The overlapping genes were investigated for overrepresented pathways and drug targets.

## 2 METHODS

### 2.1 Study Populations

We conducted the study to explore the genetic overlap between PTSD and cardiovascular outcomes, as defined by electronic health records (EHR) and heart imaging (Figure 1). Because we used previously collected, deidentified, data, this study did not require institutional review board approval. Ethics approval and participants’ consent was obtained by the original studies ^2,26–39^.

Summary statistics for GWAS of PTSD utilized in this study originated from a comprehensive meta-analysis led by Nievergelt et al. ^25^ that identified 76 autosomal loci (Supplementary Table S1). This meta-analysis encompasses findings from 88 studies gathered through the PGC-PTSD Freeze 3 data collection from three primary sources: PTSD studies employing clinician-administered or self-reported instruments (Freeze 2.5 plus subsequently collected studies, 77 studies), Million Veteran Program (MVP) release 3 GWAS utilizing the Posttraumatic Stress Disorder Checklist (PCL for DSM-IV), and 10 biobank studies incorporating EHR-derived PTSD status. In total, the study incorporated 95 GWASs, with a sample size of 1,222,882 individuals of European descent (effective sample size (Neff) = 641,533).

#### 2.1.1 GWAS of Cardiovascular and metabolic diagnoses using EHR *Million Veteran Program*

The CVD phenotype datasets utilized in this study were sourced from the MVP^40^ (see data availability), a nationwide initiative sponsored by the Department of Veterans Affairs Office of Research and Development^41^. In our investigation of the genetic overlap between PTSD and cardiovascular clinical outcomes, we incorporated summary statistics from GWAS of EHR-based phecodes. Phecodes are manually curated groups of International Classification of Diseases (ICD) codes-9/10, designed to capture clinically meaningful concepts for research purposes using the PheMap which classifies diagnoses into 17 categories (available at https://phewascatalog.org/) ^42^. We tested 141diagnoses from circulatory and 105 from endocrine/metabolic categories. A detailed description of GWAS of these traits in 458,203 individuals of European genetic ancestry in MVP is available elsewhere^40^. (Supplementary Table S2).

#### 2.1.2 UK Biobank

To replicate MVP findings, we leveraged GWAS of EHR-based phecodes data available from the UK Biobank (UKBB)^43^. Specifically, we analyzed summary statistics of GWAS of phecodes in 420,531 participants of European descent performed by the Pan-UKBB initiative ^44^. Each MVP trait was matched with a corresponding UKBB phenotype based on their phecode concordance. Because MVP phecodes 278.11, 427.21, 428.3 were not available in the UK Biobank, we paired them as follows: Phecode 278.11 (Morbid Obesity, MVP) to Phecode 278.1 (Obesity, UKBB), Phecode 427.21 (Atrial Fibrillation, MVP) to Phecode 427.2 (Atrial Fibrillation and flutter, UKBB), and Phecode 428.3 (Heart failure with reduced EF [Systolic or combined heart failure], MVP) to Phecode 428.2 (Heart failure NOS, UKBB). The details of the replication datasets are presented in Supplementary Table S3 with a comprehensive overview of clinical diagnoses, corresponding phecodes, study resources, and sample sizes.

#### 2.1.3 Other GWAS cohorts

In addition to the UKBB data, we replicated MVP findings also using 11 GWAS performed from major consortia that may include a combination of self-reported or clinical studies ^27–33^. The sample sizes of these datasets ranged from 119,715 to 1,020,441, and their diagnoses were paired with corresponding phecodes (Supplementary Table S3) ^29, 33^. For Type 2 Diabetes adjusted by BMI, we used summary statistics by the Diabetes Meta-Analysis of Trans-Ethnic association studies (DIAMANTE) Consortium ^31^, which are genetically correlated (rg) with Phecode 250.2 (Type 2 Diabetes: rg=0.992, SE=0.008; p<1×10^-300^) and Phecode 250 (Diabetes Mellitus rg=0.991, SE=0.008; p<1×10^-300^).

#### 2.1.4 GWAS of heart imaging phenotypes from UK Biobank

To also explore PTSD relationship with the structure and function of the heart and aorta, we incorporated summary-level data from a cardiac imaging GWAS ^26^. This study used information from UKBB participants who underwent comprehensive cardiovascular magnetic resonance (CMR) imaging and employed a machine learning regenerating and analyzing pipeline that resulted in 82 quantitative imaging phenotypes from 26,893 participants of European descent ^26^. These phenotypes (e.g., short-axis, long-axis, and aortic cine images) provide a detailed characterization of cardiac and aortic structure and function (Supplementary Table S2) ^26^.

#### 2.1.5 GWAS of traits used to define Life’s Essential 8

To investigate the role of other factors in the genetic overlap with CV health and PTSD, we considered the LE8 checklist defined by AHA, 2022 ^2^, which underscores eight key health factors and health behaviors related to CV well-being: eat better, be more active, quit tobacco, get healthy sleep, manage weight, control cholesterol, manage blood sugar, manage blood pressure . For our study, we identified 11 GWAS related to LE8 factors (N = 282,271 ∼ 1,320,016), including GWAS for diet (i.e., energy intake proportion of fat, carbohydrate, protein, each adjusted by body mass index, BMI) ^37^, physical activity ^36^, smoking ^35^, insomnia ^38^, BMI-adjusted waist-hip-ratio ^33^, total cholesterol ^34^, diabetes ^31^, and hypertension [MVP+UKBB]. To maximize the sample size available, we meta-analyzed hypertension GWAS (phecode 401) available from MVP and UKBB resulting in a sample size of 433,585 cases and 421,076 controls. For insomnia, we generated two GWAS datasets, one meta-analyzing GWAS of insomnia from Watanabe and colleagues ^38^ with MVP GWAS of “unable to fall asleep” (SlpFall – 188,830 cases) and the other meta-analyzing Watanabe insomnia GWAS ^38^ with MVP GWAS of “waking up in the night and not be able to fall back asleep” (SlpWakePM – 216,711 cases). To maximize the sample size available, when needed, GWAS datasets were meta-analyzed using a fixed-effects inverse variance-weighted model available in METAL ^45^. A detailed description of all cohorts is presented in Supplementary Table S2.

### 2.2 Proteomic cohorts

#### 2.2.1 Brain Proteomic Datasets

The Religious Orders Study and Rush Memory and Aging Project (ROS/MAP) proteome expression weights were generated by Wingo et al. ^46^, using dorsolateral prefrontal cortex (dlPFC) tissues available from 376 individuals of European ancestry^46^. Briefly, their analyses included calculating the normalized abundance of 8,356 proteins. Among these, 1,475 proteins exhibited significant cis associations with genetic variation. The weights assigned to these proteins were used in our proteome-wide association study (PWAS). Comprehensive details regarding sample description, proteomic analysis, quality control, and statistical analyses can be found in the original paper by Wingo et al ^46^. Additionally, we utilized dlPFC genetic-proteomic data from the Banner Sun Health Research Institute (BANNER). This dataset comprises brain dlPFC samples from 152 individuals of European descent, with 8,168 proteins included in proteomic profiles following quality control ^46^. Through the integration of proteomic data and SNP genotypes, 1,139 proteins exhibited significant heritability to genetic variation. Therefore, ROS/MAP (N=376) and BANNER (N=152), encompass a total of 1,797 proteins. (See Data Availability and Links).

#### 2.2.2 Atherosclerosis Risk in Communities (ARIC) Study

To study the plasma proteome, we utilized data from Zhang and colleagues ^47^. Proteomic data was derived from summary-level information obtained from The Atherosclerosis Risk in Communities (ARIC) Study, focusing on 7,213 individuals of European descent ^47^. The study measures plasma proteome levels using the SOMAmer-V4 platform. After quality control, 1,348 plasma proteins exhibiting associations with common variants in cis regions were investigated in our PWAS study (See Data Availability and links).

#### 2.2.3 UKBB-Plasma Proteome Project (UKB-PPP)

In addition to the ARIC Study, we tested data from the UK Biobank Pharma Proteomics Project (UKB-PPP). This project provides the plasma proteomic profiles of 54,219 participants along with genome-wide genotypes, exome sequencing, whole-body magnetic resonance imaging, electronic health records, blood and urine biomarkers, as well as physical and anthropometric measurements. UKB-PPP used Olink proteomics assay for proteome assessment, and reported expression weights for protein quantitative trait loci (pQTLs) for 2,923 unique proteins ^48^.

### 2.3 Risk loci from PTSD GWAS

To identify genomic loci of interest in PTSD GWAS summary statistics, as per original study we used Functional Mapping and Annotation of Genome-Wide Association Studies (FUMA)^49^. Considering LD weights from the European reference populations of the 1000 Genomes Project, resulting in 76 independent genomic risk loci (P < 5×10⁻^8^, r^2^<0.6, MAF≥0.01, LD distance <250kb) (Supplementary Table S1) as reported by Nievergelt et. al 2024.

### 2.4 Shared regions and variants between PTSD and the CVD traits

#### Local Analysis of [co]Variant Association (LAVA)

LAVA ^50^ was employed to identify loci exhibiting local genetic correlation between PTSD and a total of 328 CV traits across three categories . We applied univariate LAVA models at the 76 PTSD risk loci. Loci that reached false discovery rate significance at 5% (FDR q<0.05) for the heritability univariate models were selected for local genetic correlation between PTSD and each one of the CV/LE8 traits. Bivariate LAVA results were filtered to include only those cases where both traits exhibited significant univariate heritability on the locus, as determined by an FDR q < 0.05. Sample overlap was estimated with LD score regression with the European LD scores calculated from the 1000 Genomes as reference (https://github.com/bulik/ldsc). Variants that were not SNPs (e.g., indels) and SNPs that were strand-ambiguous, multi-allelic, and had a minor allele frequency (MAF) <0.01 were excluded.

#### Colocalization analysis

Colocalization is a statistical approach used to assess whether pairs of traits share a putative causal variant within the same genomic region. COLOC employs a Bayesian approach, considering various variant-level hypotheses and calculating Bayes factors from SNP effect estimates and standard errors ^51^. The variant-level hypotheses correspond to: H0 (no association to either trait), H1 (association to only trait 1), H2 (association to only trait 2), H3 (associations to both traits with different causal variants), and H4 (associations to both traits with shared causal variants) ^52^. In our study, each genomic region was expanded by 500kb on either side of the position. . COLOC ^51^was applied to assess colocalization between PTSD and the significantly correlated trait. Results with H4 or H3 hypothesis probabilities ≥80% were considered as shared. For replication, we used data from UKBB and other cohorts considering H4 or H3 hypothesis probabilities ≥70% as strong evidence of shared variants.

#### Tissue and molecular-profile-based prioritization of genes within colocalized regions

We used the UCSD Genome Browser Tool (see data availability and links) to identify genes within the colocalized regions with significant H4/H3 probability for PTSD with at least one CVD trait. Then, we gathered colocalization probabilities (H3 and H4) for molecular profiles of those genes and all reported phenotypes from published GWAS studies using OpenTargets ^53^. By manually examining the traits, we defined the traits as psychiatric and/or CVDs.

### 2.5 Observational Analysis in All of Us Research Program

Started in 2018, the National Institutes of Health’s All of US (AoU) Research Program has enrolled over 700,000 diverse participants, emphasizing health equity in the context of precision medicine. Leveraging EHR data available for 254,700 AoU participants ^54^. Phecodes were derived from mapping International Classification of Disease ICD9/ICD10(CM) codes using Phecode Map (see data availability and links). Logistic regression using the PheWAS R package was employed to assess the association of PTSD (13,877 cases) with CV phecodes identified in MVP and UKBB analyses, considering three models (i) base-model (sex, age, self-reported race), (ii) socioeconomic (SES)-model (bases model covariates plus BMI, smoking, deprivation index), and (iii) depression-model (SES-model covariates plus depression diagnosis: phecode 296.2-Ncases:72,143).The self-reported demographic characteristics of AoU are as follows: females (61.8%), average age in years = 54 ±17 (mean±SD), 57.17% White, 19.4% (self-identified race), Black or African American, 3.54% Asian, 0.62% Middle Eastern/North African, 1.92% More than one population, 0.11% Native Hawaiian/Pacific Islander, and 17.1% None of the above), while 44.1% were smokers (past and current).

### 2.6 Proteome-Wide Association Study

Proteome-wide associations of PTSD and its colocalized traits were estimated We integrated GWAS data with tissue-specific pQTLs. For the colocalized phecodes, PWAS was conducted using the meta-analysed GWAS combining MVP and UKBB as described above, using the FUnctional Summary-based ImputatiON (FUSION) (available at http://gusevlab.org/projects/fusion/)^55^.

PWAS using weights from UKB-PPP plasma proteomewas performed using Summary-based Mendelian Randomization (SMR) ^56, 57^. The analysis was performed using SMR default settings (https://yanglab.westlake.edu.cn/software/smr/#Overview) ^56^. The results were adjusted for multiple testing with FDR correction (FDR q<0.05).

### 2.7 In-silico functional Analysis

#### 2.7.1 Pathway enrichment

To gain insight into the shared mechanisms between PTSD and CVD traits, we conducted pathway enrichment analysis based on the genes detected in brain and blood proteome-wide analyses. Pathway enrichments were sought using a web-based tool, g:Profiler (https://biit.cs.ut.ee/gprofiler/gost) among three libraries: Gene Ontology (GO), Kyoto Encyclopedia of Genes and Genomes (KEGG), and Reactome Pathway Database (REAC) ^58^. GO Database contains three functional domains: cellular component (CC), molecular function (MF), and biological process (BP). A significance threshold of FDR q<0.05 was used to identify significant pathways.

#### 2.7.2 Drug repurposing in research context

Significant genes from the brain and plasma PWAS and the prioritized genes from the colocalization analysis were used for identifying drugs from the drug-gene interaction database (DGidb)^59^. Secondly, we obtained information including drug ID, clinical uses and adverse effects from stored at OpenTargets (https://www.opentargets.org/) which are originally sourced from FDA database. The OpenTarget results were compared with current CVD and PTSD treatments, to identify drugs with overlapping functions and suggest new potential therapies.

## 3 Results

### 3.1 Genetically correlated loci between PTSD and CVD Traits

We investigated 76 genome-wide significant (GWS) risk loci associated with the PTSD GWAS (P < 5×10⁻⁸, Supplementary Table S1) for local genetic correlation with GWAS of 246 CV-related phecodes from MVP, 82 cardiac imaging traits from UKBB, and LE8-related phenotypes from various studies. We observed statistically significant SNP-based heritability at 73 loci for all the phenotypes investigated (Supplementary Table S4). For CV-phecodes, we identified 112 local genetic correlations with PTSD across 33 loci (FDR q<0.05; Figure 2, Supplementary Table S5). Among these, 67 were related to circulatory system, and 45 to endocrine/metabolic phecodes. . Additionally, four PTSD-associated loci presented local genetic correlations between PTSD and 14 heart imaging traits (FDR q<0.05; Figure 2, Supplementary Table S5). For LE8-factors, 92 local genetic correlations with PTSD were identified across 44 loci (Figure 2, Supplementary Table S5). Notably, fat dietary intake was the only LE8-trait that was not genetically correlated with PTSD at any of the investigated loci. Overall, most local genetic correlations were positive. Among the few negative local genetic correlations, four were between PTSD and total cholesterol across different loci and six at 15q26.1 between PTSD and multiple phenotypes (Figure 2, Supplementary Table S5).

**Figure 2:**
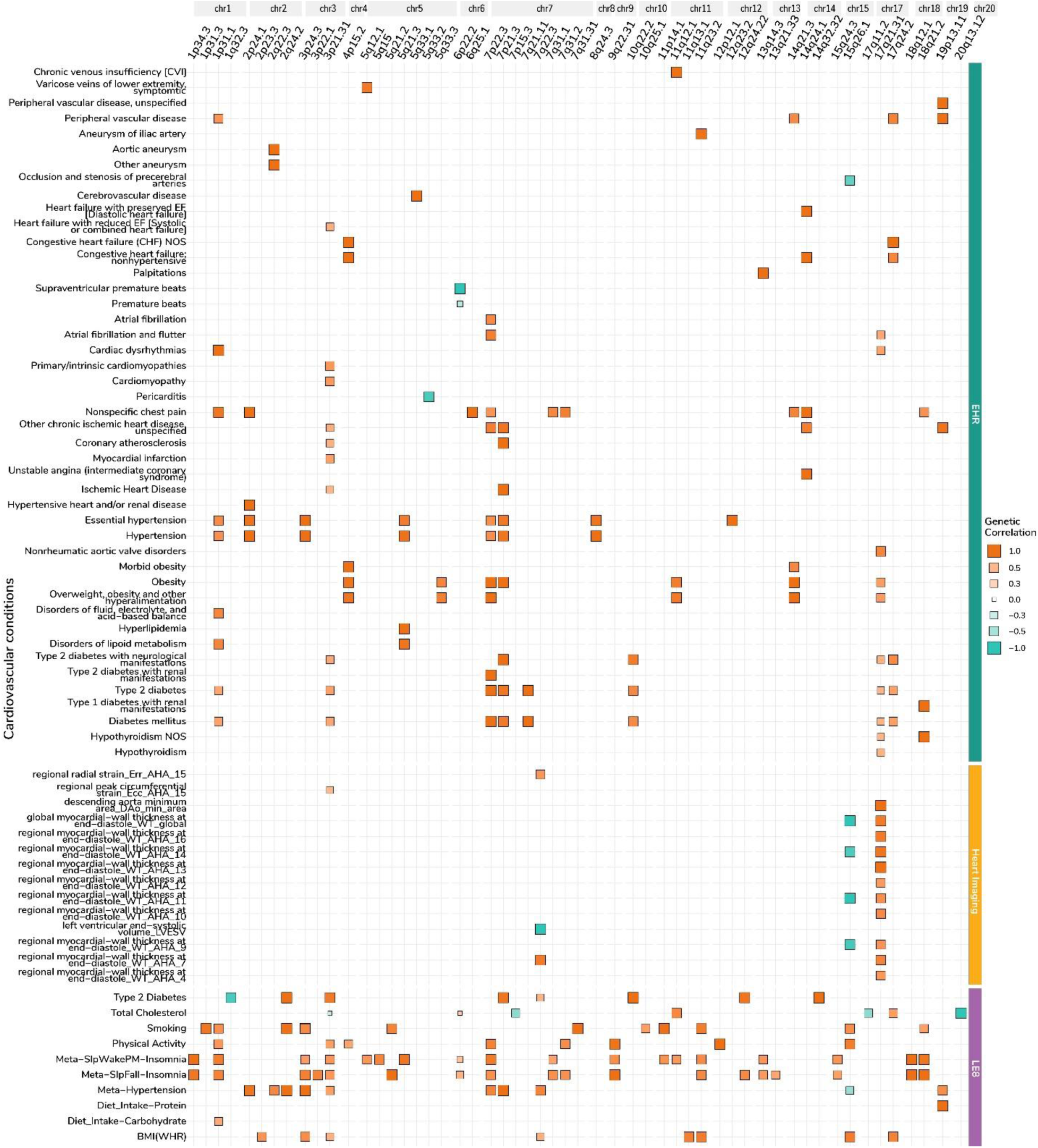
Local Genetic Correlation between PTSD and CV conditions: Matrix plot of local genetic correlation between PTSD and conditions grouped by their CV category (left y-axis); the EHR-based CV definitions (i.e., phecodes) are from Million Veteran Program. The x-axis shows loci as cytobands grouped by chromosomes. The positive correlation is denoted in orange, cyan indicates negative correlation, and the size of the squares corresponds to the magnitude of the genetic correlation.

Considering locus-specific results, the 17q21.31 region exhibited the highest number of genetically correlated traits (N=21; Figure 2, Supplementary Table S5), including 10 CV-related phecodes (e.g., cardiac dysrhythmias, nonrheumatic aortic valve disorders, diabetes mellitus, overweight, and hypothyroidism) and 11 heart imaging traits (e.g., various sections of myocardial-wall thickness at end-diastole and ascending aorta minimum area).

### 3.2 Shared causal variants between PTSD and CV conditions within genetically correlated loci

We investigated whether the local genetic correlation between PTSD and CV-related phenotypes is due to a shared causal variant or causal variants in linkage disequilibrium (colocalization hypotheses H4 and H3, respectively) ^52^. Our colocalization investigation identified 20 CV-related phecodes that shared the same causal SNP with PTSD across 11 loci (H4-PP ≥ 80%, Figure 3A; Supplementary Table S6) and 19 CV diagnoses for which causal variants were in LD with PTSD variants (H3-PP ≥ 80%, Figure 3A; Supplementary Table S6). To replicate the colocalization findings observed using MVP phecodes, we repeated the analysis using UKBB phecodes and other cohorts which incorporate a combination of EHR and self-report definitions. Specifically, we matched 24 CV phecodes in UK Biobank (N=420,531) ^44^ and 11 GWASs from major consortia comprising of self-reported and/or clinical data (N=∼1,320,016) ^27–33^. In UKBB, 13 out of the 24 examined traits displayed replications (H4/H3-PP ≥70%), revealing consistent colocalization patterns with PTSD across 8 loci (Supplementary Table S6). In the other cohorts, validation was achieved for 5 phenotypes (i.e., myocardial infarction, coronary artery disease, atrial fibrillation, type 2 diabetes, and BMI) across 8 distinct loci (H4/H3-PP ≥70%; Supplementary Table S6). Across discovery (in MVP) and replication (in UKBB and other cohorts), obesity and being overweight demonstrated the highest posterior probability of sharing the same causal variant with PTSD in locus 4p15.2 (H4-PP=1, a shared causal SNP - rs34811474 - *ANAPC4*).

**Figure 3:**
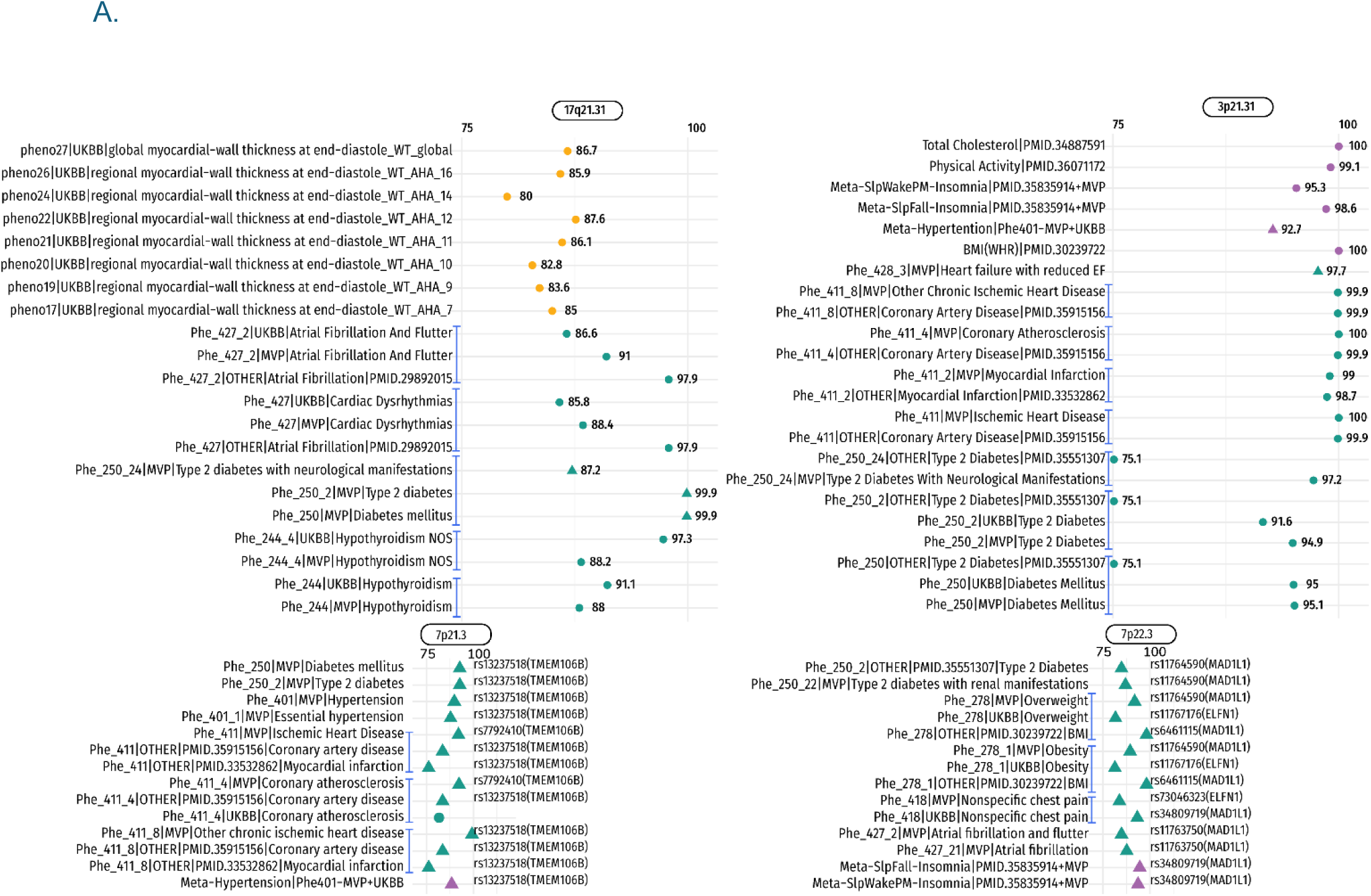
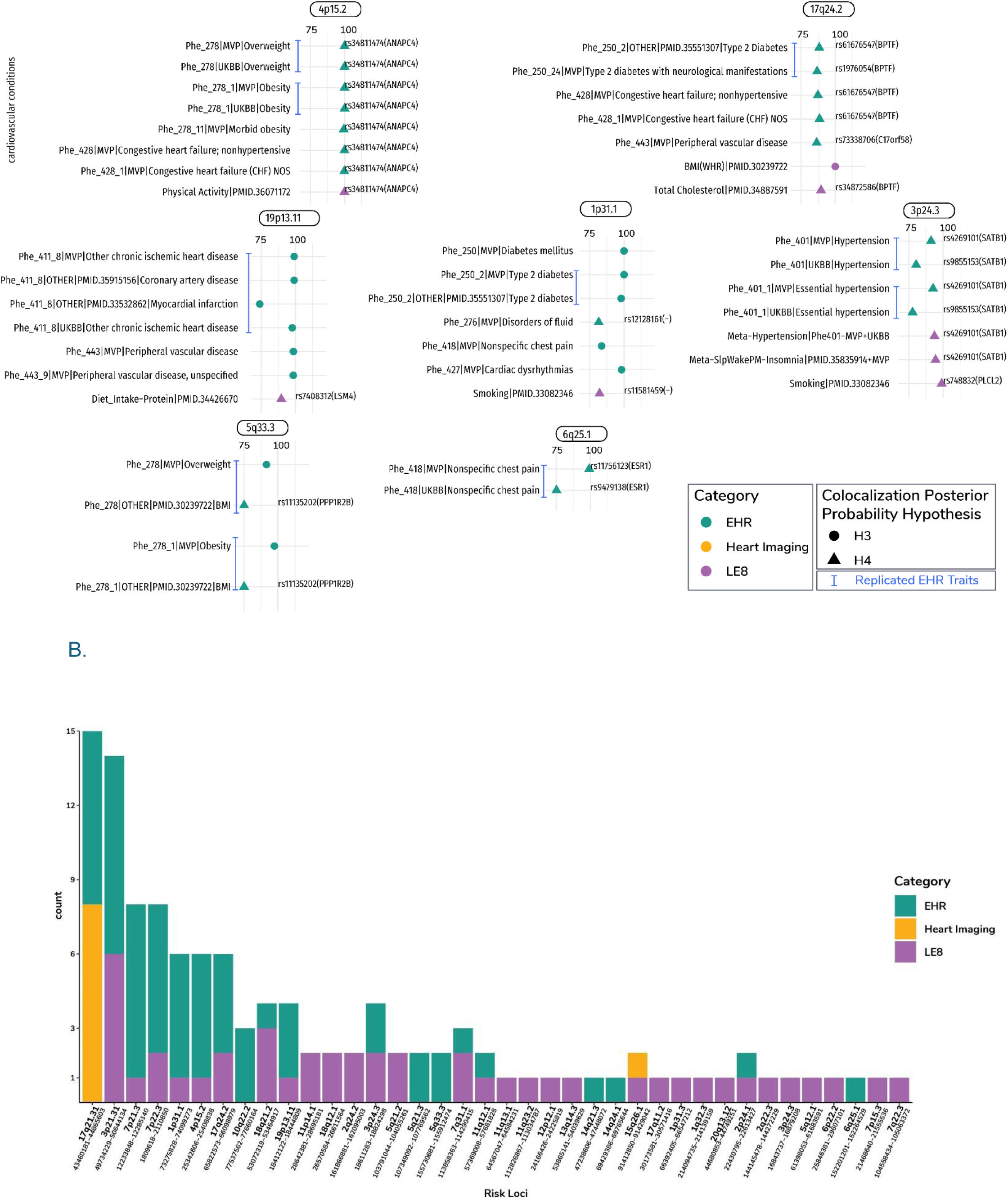
Shared causal variants between PTSD and CV conditions: A) *Top*. The x-axis shows colocalization probability(different LD-linked causal variants-H3 hypothesis; same causal variant – H4 hypothesis), and y-axis is marked with CV conditions. between PTSD and CV conditions based on colocalization. A subset of 11 loci that replicated for EHR-derived diagnosis are indicated with a blue line. The shared causal variants based on H4 hypothesis are labelled and their nearest genes are in parenthesis. We highlight genes at each locus that had colocalization evidence with molecular profiles (gene/splicing/proteome expression) and CV conditions (Supplementary Table S7); B) *Bottom*. A stacked bar plot showing number of traits (y-axis) observed at each locus, marked as cytoband and position range in base pairs (x-axis). The traits are grouped into three CV categories.

Among the heart imaging traits, there was colocalization on two loci for 8 traits. Within locus 17q21.31, PTSD-variants were in LD with causal variants related to seven different levels of measurements of regional myocardial-wall thickness at end-diastole, and the global myocardial-wall thickness at end-diastole (H3-PP≥80.0%). In locus 15q21.31, PTSD and the global myocardial-wall thickness at end-diastole share the same causal trait (H4-PP=92.5%, causal SNP - rs17514846- *FURIN*).

Considering LE8 checklist, we observed statistically significant PTSD colocalization (H4/H3 ≥80%) in 31 loci (Figure 3A; Supplementary Table S6; Supplementary Figure S1). While no colocalization was observed between PTSD and dietary carbohydrate intake, the other LE8-related phenotypes shared the same PTSD casual SNP in 20 loci collectively (H4-PP ≥80%; closest gene (shared causal variant), *ANAPC4* (rs34811474), *ARHGAP15* (rs10191758), *BPTF* (rs34872586), *CDH2* (rs7243332), *FOXP2* (rs1476535, rs8180817), *IP6K1* (rs11130221), *KMT2E* (rs2470937), *LSM4* (rs7408312), *MAD1L1* (rs34809719), *NCAM1* (rs7106434), *NCOA5* (rs6032660), *PDE4B* (rs2310819), *PLCL2* (rs748832), *PROX1* (rs340874), *SATB1*(rs4269101), *TANK* (rs197261), *TMEM106B* (rs13237518), non-coding regions (rs11581459, rs325500, rs4275621)) Supplementary Table S6; Supplementary Figure S1). Interestingly, physical activity shared the same causal variant with PTSD in locus 4p15.2 (H4-PP=99%) that we observed with respect to obesity and being overweight (shared causal SNP - rs34811474 - *ANAPC4*). Both total cholesterol and BMI–adjusted-waist-to-hip ratio had the highest posterior probability of having causal variants in LD with PTSD-associated SNPs in 6p22.2 and 3p21.31, respectively (H3-PP=100%, Supplementary Table S6; Supplementary Figure S1).

Overall, 17q21.31 and 3p21.31 regions exhibited the highest number of CV-related phenotypes (Figure 3B). Specifically, 17q21.31 showed colocalization of PTSD with seven CV phecodes and eight heart imaging phenotypes, while 3p21.31 colocalized PTSD with eight CV phecodes and six phenotypes related to LE8 checklist.

To prioritize genes within the colocalized regions, we first identified 506 genes physically located in the 38 loci that showed evidence of colocalization between PTSD and CV conditions. We leveraged multi-tissue molecular profiles (gene, proteome, and splicing expression) and CV traits available from OpenTargets platform ^51, 53^. We identified 270 genes that had H4 or H3 hypothesis probability ≥0.6 of colocalizing with 201 different CV conditions across 124 tissues. Among the highest H4 associations (H4-PP=100%), there were multiple phenotypes related to the body electrical impedance, BMI, blood lipids, blood pressure, hemoglobin A1c in multiple tissues, smoking, sleep duration, and hypothyroidism. (Supplementary Table S7).

### 3.3 Observational association of PTSD with CV-related diagnoses in All Of US cohort

To further investigate the comorbidity between PTSD, and 13 CV diagnoses observed in our genetically informed analysis, we conducted an observational analysis using EHR data for circulatory and metabolic diagnses from AoU cohort *(as per phecodes: 244-hypothyroidism, 244.4-hypothyroidism NOS, 250-diabetes mellitus, 250.2-Type 2 diabetes, 278-Overweight, obesity, and hyperalimentation, 278.1-obesity, 401-hypertension, 401.1-essential hypertension, 411.8-other chronic ischemic heart disease, 418-nonspecific chest pain, 427-Cardiac dysrhythmias, 427.2 atrial fibrillation and flutter, and 411.4-coronary atherosclerosis)*).

Specifically, we tested the association of PTSD (13,877 cases) with 13 CV diagnoses (Supplementary Table S8) considering three adjustment models: i) base-model (covariates:-age, sex, and self-reported race); ii) SES-model (base-model covariates and deprivation index, smoking, and BMI), iii) depression-model (SES-model and depression diagnosis-[phecode 296.2]). PTSD was significantly associated with all 13 CV phecodes across all three models (p<5.15×10^-6^; Supplementary Table S8). However, while there was no difference between the estimates obtained from base and SES-models, we observed a reduction in effect sizes observed in the depression model compared to the base model ranging from 84% for chronic ischemic heart disease (odds ratio, OR= 2.61 vs 1.41, p-difference=5.44×10^-19^) to 48% for hypothyroidism (OR= 1.78 vs 1.2, p-difference=5.7×10^-25^). Nevertheless, the effect sizes observed accounting for depression comorbidity confirm the relationship linking PTSD to CV-related traits.

### 3.4 Tissue Enrichment for PTSD, CV diagnoses, heart imaging and LE8 traits

To gain biological insights into PTSD and CV conditions, we further employed in-silico genetic approaches to identify tissues that might overlap due to similar gene expression profiles. We limited our analyses to CV traits that showed multiple levels of evidence for genetic overlap: 13 CV diagnoses (meta-analyzed between UKBB and MVP), 8 heart imaging traits and eight LE8 traits.

Tissue enrichment test systematically models tissue-specific gene expression data and GWAS of trait, prioritizing disease specific causal tissues. We identified 6 tissues that are enriched based on gene expression in PTSD: the limbic system (p=4x10^-6^), the cerebral cortex (p=6x10^-5^), the brain (p=4x10^-6^), the entorhinal cortex (p=4x10^-4^), the hippocampus (p=10^-3^), and the brain cortex (p=9x10^-4^). These tissues were not FDR significant in other traits, but were nominally significant for nonspecific chest pain, overweight, ‘overweight, obesity and other hyperalimentation’, physical activity, smoking, diet intake proportion of protein, and insomnia (SlpFall-difficulty in falling asleep, and SlpWakePM-difficulty in falling asleep after waking up in the middle of the night) (p<0.05; Supplementary table S10; Supplementary Figure S2). No heart imaging trait remained significant in the multi-tissue enrichment analysis.

### 3.5 Proteome-Wide Association Study (PWAS): integrating genetic variants from GWAS and proteome expression in brain and blood tissues

PWAS studies combine effect-estimate of genetic variants on diseases, and abundance or expression of proteins, thereby prioritizing genes that may be associated with disease/traits via altered proteome expression. We tested gene-associations leveraging genetically regulated proteome expression in dlPFC and blood with respect to PTSD and phenotypes highlighted by local genetic correlation and colocalization analyses. This included 13 CV phecodes, seven heart imaging traits, and LE8 factors. To maximize statistical power, we meta-analyzed GWAS of each of the 13 CV diagnoses from MVP and UKBB to improve statistical power for gaining insights into overlapping mechanistic pathways [N total = 865,527]. Details regarding each meta-analyzed CV diagnosis are available in Supplementary Table S9. Leveraging weights derived from dlPFC-specific pQTLs, we identified 122 genes associated with both PTSD and CVD phenotypes (FDR q<0.05; Figure 4, Supplementary Table S11). The majority of these PTSD associations were shared with LE8 factors (N=109). Several genes demonstrated proteomic associations across CV phecodes, heart imaging phenotypes, and LE8 factors (e.g., *ATG7*, *CCDC92*, *CNNM2*, *DNM1*, *FAM134A*, *SIRPA*, *SNX32*, and *TR0IM47*). Other pleiotropic genes included *CCDC92*, *SIRPA* and *LRRC37A2* that were associated with PTSD and 20 or more CV-related phenotypes (Supplementary Table S11). The strongest proteome-wide association with PTSD was observed with *ICA1L* (Z=6.89, p=5×10^-12^) that was also associated with several CV phecodes such as atrial fibrillation (Z=2.9, p=4×10^-59^), coronary atherosclerosis (Z=16.2, p=5×10^-50^), and T2D [EHR-MVP+UKBB] (Z=3.87, p=1.07x10^-4^). While these associations were positively related to increased *ICA1L* proteomic expression, we also observed an inverse relationship with total cholesterol (Z=-18.86, p=2×10^-79^), insomnia (Z=-6.03, p=2×10^-9^), physical activity (Z=-3.41, p=6×10^-4^), and smoking (Z=-4.85, p=10^-6^). The strongest inverse association with PTSD was *KHK* (Z=-6.86, p=7×10^-12^), which was also negatively associated with several other phenotypes, such as T2D (Z=-5.22, p=2×10^-7^), hypertension (Z=-4.59, p=4×10^-6^), unspecified chronic ischemic heart disease (Z=-4.04, p=5×10^-5^), hypothyroidism (Z=-3.94, p=8×10^-5^) protein intake (Z=-3.84, p=10^-4^), coronary atherosclerosis (Z=-3.82, p=10^-4^), and nonspecific chest pain (Z=-3.32, p=9×10^-4^). *KHK* was positively associated with BMI (Z=3.49, p=5×10^-4^) and LE8-T2D (Z=3.56, p=4×10^-4^). Total cholesterol demonstrated the highest number of overlapping genes with PTSD, with 56 genes overlapping in the brain proteome including genes with the highest effect estimate such as *PLCG*1 (Z = -18.349, p=3.37×10^-75^).

**Figure 4:**
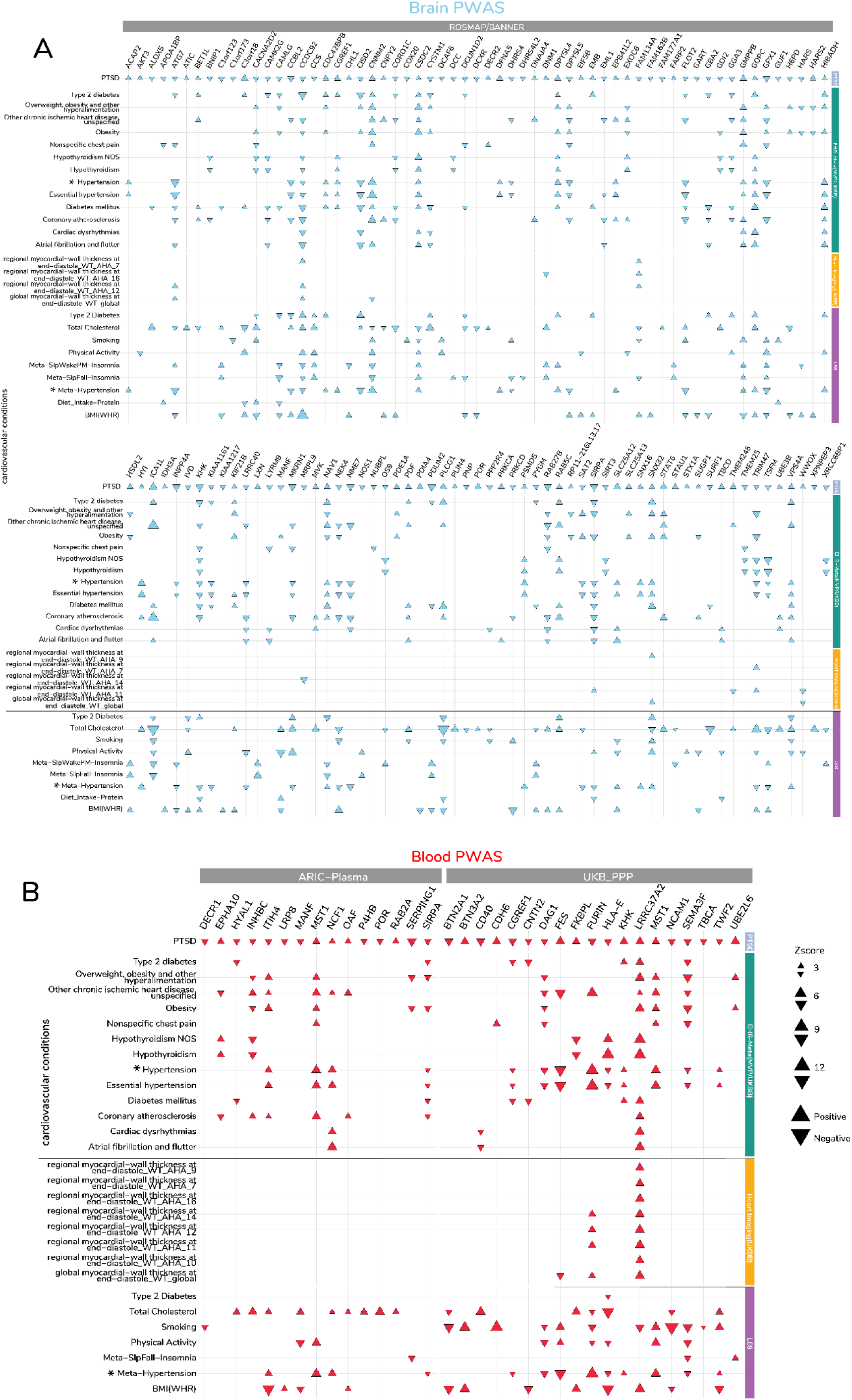
Shared genes and pathways between PTSD and CV conditions based on proteome-wide associations: Distribution of z-scores across significant PWAS genes between PTSD and CV conditions using A) brain proteome in blue and B) blood proteome in red. Genes are grouped based on two blood-based proteome panels/brain-based panel (y-axis) and respective CV conditions (x-axis). Significant genes are shown as red (blood) or blue (brain) triangles, wherein triangles facing up and down represent positive and negative z-scores, respectively.

Leveraging blood-proteome expression from two different studies (ARIC, and UKBB-PPP), we identified 33 genes associated with both PTSD and at least one or more of the 13 CV diagnoses, 8 heart imaging traits, and LE8 factors investigated (Supplementary Table S11). The strongest positive association with PTSD was *FES* (Z=6.15, p=8×10^-10^), which was also positively associated with smoking (Z=5.95, p=3×10^-9^) and physical activity (Z=4.19, p=3×10^-5^). *FES* proteomic expression exhibited negative associations with essential hypertension (Z=-9.92, p=3×10^-23^), unspecified chronic ischemic myocardial disease (Z=-8.51, p=2×10^-17^), and global myocardial wall-thickness at end-diastole (Z=-4.08, p=5×10^-5^). The strongest negative association with PTSD was observed with *CD40* (Z=-5.27, p=10^-7^), which was also negatively associated with atrial fibrillation and flutter (Z=-3.73, p=2×10^-4^) and positively associated with total cholesterol (Z=7.17, p=8×10^-13^). Considering both blood and dlPFC, *SIRPA*, *MANF*, and *POR* exhibited cross-tissue proteome-wide associations with PTSD and CVD traits (Supplementary Table S11).

By comparing genes prioritized from colocalized regions and genes identified from PWAS, we identified 403 distinct genes with 17 overlapping between the two methods: *BTN2A1, BTN3A2, C3orf18, CACNA2D2, CD40, DAG1, FES, FURIN, GMPPB, GPX1, HYAL1, LRRC37A2, MST1, NCAM1, SEMA3F, SERPING1, UBE2L6*.

### 3.6 Pathway Enrichment

Considering genes identified by the tissue-specific PWAS, we identified 25 pathways overrepresented by the PTSD&CV proteome-wide significant genes in the dlPFC and 36 pathways in the blood (Supplementary Table S12). Among the dlPFC PWAS genes, in addition to basic cellular functions (Supplementary Figure S3; Supplementary Table S12), we observed metabolic and calcium modulating pathways: “oxidoreductase activity, acting on the CH-OH group of donors, NAD or NADP as acceptor” (FDR P-value = 3.92×10^⁻2^), “Calmodulin-induced events” (FDR P-value =3.97×10^⁻2^), the “CaM pathway” (FDR P-value =3.97×10^⁻2^), and “Ca-dependent events” (FDR P-value =4.95×10^⁻2^). Among the pathways overrepresented by the plasma PWAS genes, in addition to biological and cellular processes (Supplementary Figure S4; Supplementary Table S12), we observed several immune and neuronal processes Such as “response to stimulus” (FDR P-value= 5.84×10^⁻4^), “regulation of immune response” (FDR P-value =4.98×10^⁻3^), “regulation of immune system process” (FDR P-value =7.33×10^⁻3^), “regulation of response to stimulus” (FDR P-value= 0.011), neuron projection development (FDR P-value= 0.014).

### 3.7 Drug repurposing in research context

To contextualize the role of reported shared genes between PTSD and CV conditions, we performed a prototype approach. We identified drugs that target these shared genes. We then analyzed if any of these drugs have common gene targets, and categorized their adverse effects into cardiovascular or psychiatric side effects. This approach helps us see how these shared genes might influence both types of conditions and the potential risks and benefits of the drugs involved.

We identified 74 approved drugs targeting 30 genes that either designated for psychiatric (as a range of psychiatric drugs can be used for treating PTSD and associated symptoms) or cardiovascular conditions (Supplementary Table S13, Figure 5). Looking at gene target that overlap between psychiatric and CV conditions, we found *DRD2*, *GFAP*, *POR* and *NOS1* to be targets of several CV conditions including hypo/hypertension, diabetes, cholesterol and heart failure. Only Prazosin, an alpha-1 adrenergic receptor antagonist was the only drug that is known to treat hypertension, and has off-label benefit for PTSD-associated nightmares^60, 61^.

**Figure 5:**
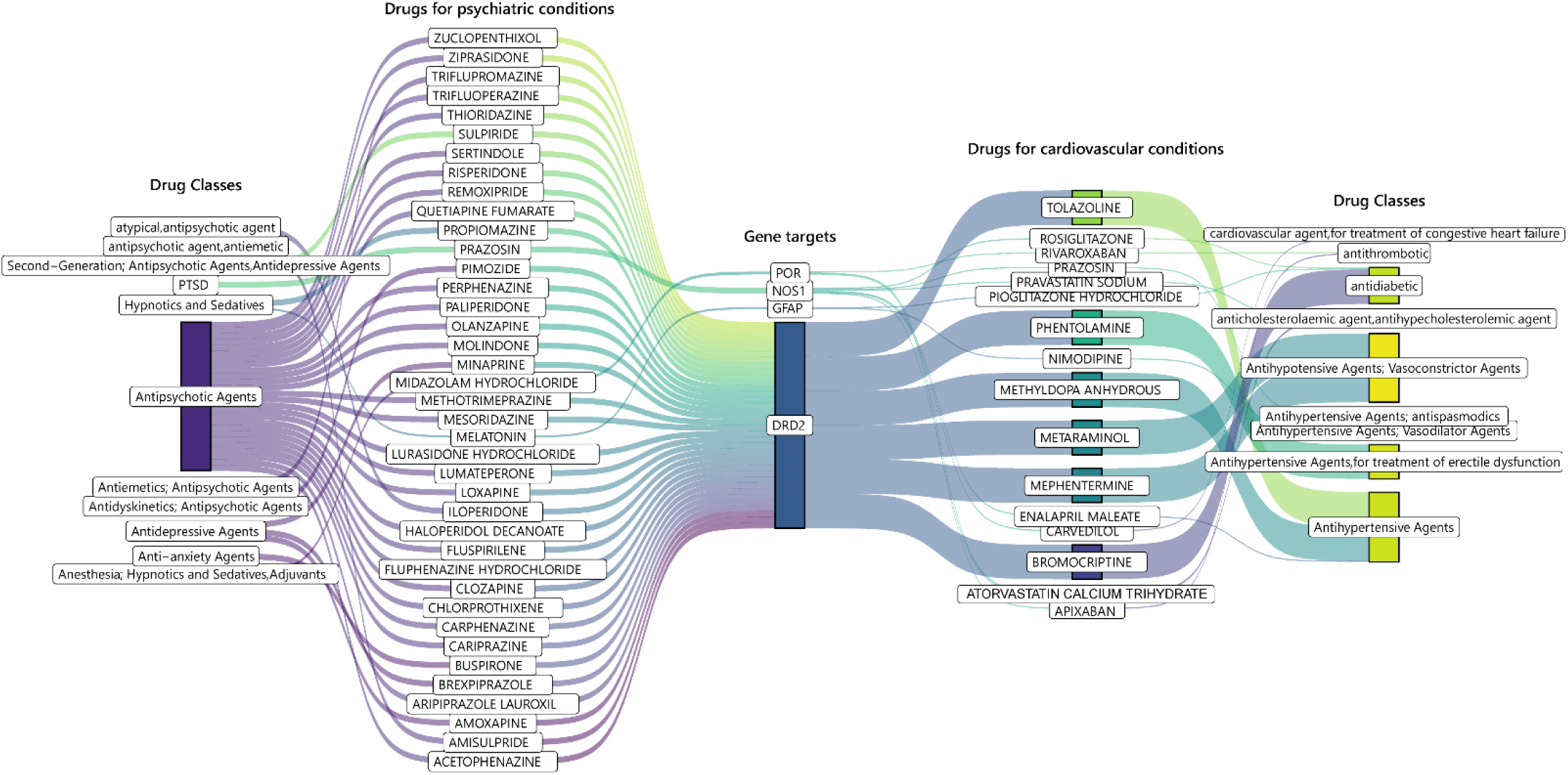
Comparing common drugs and their gene-targets between PTSD and CV conditions: This Sankey plot shows the psychiatric drugs and their classes (first & second panel) that target genes – *DRD2, GFAP,POR* and *NOS1* (third panel), which are also targeted by CV drugs (third panel) and their corresponding CV categories (fourth & fifth panel) (see Supplementary Tables for more details).

We also investigated adverse effects of these drugs, and among the 45 psychiatric drugs most cardiovascular-system-related adverse effects included increased weight or weight fluctuations, tachycardia and QT prolongation, which indicates a disturbance in heart ventricle chamber signal transmission ^62^. Other CV adverse effects of the psychiatric drugs include blood glucose increase, diabetes, orthostatic hypotension, and dyslipidemia.

Conversely, among the 30 CVD drugs, adverse psychiatric effects were less observed, and included depression, suicide attempt, cognitive disorder, and hallucination. (Supplementary Table S14).

## 4 Discussion

To our knowledge, we provide the first comprehensive study assessing the genetic overlap between PTSD and CV phenotypes in more than 1 million individuals, integrating multi-omics information with EHRs, functional and structural cardiac imaging measurements, and CV health factors. We report causal variants for CV diagnoses and imaging traits that are shared with PTSD, while CV health behaviors as per AHA’s LE8 show a broader genetic overlap across several loci. Among the EHR CV traits, colocalization analyses of discovery and replication cohorts indicated loci 3p21.31, 17q21.31, 7p22.3, and 7p21.3 as potential PTSD-CVD pleiotropy hotspots. Specifically, loci 3p21.31 and 17q21.31 exhibited different LD-linked causal variants, while loci 7p22.3 (*MADIL1, ELFN1*) and 7p21.3 (*TMEM106B, VWDE, THSD7A*) had the greatest number of causal variants shared between PTSD and multiple CV conditions (e.g., hypertension, diabetes, coronary artery disease, obesity/overweight, atrial fibrillation, and non-specific chest pain). Although some of these loci have been identified in the context of psychiatric disorders ^63–65^, we additionally observed genes at these loci also colocalize between multi-tissue molecular profiles of several cardiovascular traits, highlighting their significant role as a potential common denominator to PTSD and CV conditions. Often genetic data is proxy for diagnostic risk, and mirrors association, but is limited in adjusting for additional confounders without losing statistical power. We confirmed the specificity of PTSD-CV genetic relationships using epidemiological observation from EHR data of 249,906 AllofUS participants. We observed that PTSD diagnosis is associated with 13 CV diagnoses (identified using genetic data from MVP EHR data and replicated in UKBB EHRs), even after accounting for smoking, BMI, deprivation index, and depression diagnosis. Interestingly, 4 of the 13 diagnoses overlap with CV health factors from the LE8 traits – type 2 diabetes, obesity, overweight, and hypertension, which is parallel to genetic study of ideal cardiovascular health based on LE 7 ^66^.

To identify potentially actionable gene targets that overlap between PTSD and CV conditions, we leveraged the cumulative effect of genetic variants on protein expression. We identified twice as many genes using brain-based proteome expression weights than with blood. We also observed interesting relationships, such as total cholesterol demonstrating negative genetic correlation with PTSD at multiple loci, and almost half of the gene associations have opposite effect estimates between PTSD and total cholesterol. These observations may contribute to the abnormal total cholesterol reported in individuals with PTSD ^67^. Additionally, we report many shared proteomic associations of PTSD and CVD to further identify underlying common disease mechanisms and therapeutic targets. Among them, *LRRC37A2* was related to the most CVD traits (N=20), followed by *MST1* (N=18). *LRRC37A2* has been associated with coronary artery disease ^68^, cardiorespiratory fitness ^69^, and thyroid function ^70^ . *MST1* activation has been linked to the pathogenesis of cardiovascular and metabolic diseases ^71^ while its downregulation in the hippocampus appears to be protective in the context of stress-related mental health conditions (e.g. PTSD) ^72^. *NCAM1* showed the most statistically significant proteomic association with smoking. This is in line with previous evidence linking this locus to smoking ^73^. Other PTSD-CV shared genes with supporting evidence from previous CV studies included *TMEM106B* ^74–76^ , *MAD1L*1 ^77, 78^, *SATB1* ^79^, *PLCL2* ^80^, *FURIN* ^81, 82^, *FOXP2* ^83^*, ESR1* ^84^, and *CNNM2* ^85^. Pathway enrichment analyses highlighted different blood vs. brain patterns. Specifically, genes identified by the blood-based PWAS were enriched for immunological and neuronal processes, while the brain-based PWAS genes were related to carbohydrate metabolism and calcium-modulating functions. These findings converge with previous hypotheses related to the role of immune-metabolic signaling ^86, 87^ and calcium dysregulation ^88–92^ in PTSD and CV pathogeneses.

The drug repurposing analyses identified *DRD2, GFAP, NOS1 and POR* as common gene targets of several PTSD and CV drugs. Prazosin was common to PTSD and CV conditions, although with mixed results for alleviation of PTSD and related symptoms. Altered levels of *GFAP* expression are associated with PTSD^93^, and thrombotic injury to the vascular muscle leads to secretion of GFAP^94^. Furthermore, the drugs used for PTSD treatment we identified in our drug-gene-interaction analysis have reported adverse CV effects such as weight gain, weight changes, cardiac arrythmia, abnormal blood pressure, and diabetes^95, 96^. *NCAM1* that colocalizes with BMI, is on the same locus as *DRD2* at 11q23.2, with both genes having molecular colocalization with insulin-like growth factor, blood pressure, and BMI. On the other hand, CVD drugs that exhibit psychiatric adverse effects such as depression, and cognitive dysfunction which have been associated with serotonergic ^97^, and antidepressant response ^98–100^ respectively. These findings suggest a common role of the drugs in the two different diseases, where multiple mechanisms may be involved, either protective or detrimental.

While we provide a comprehensive assessment of the biology shared between PTSD and several CV conditions, our study has some limitations. Due to the well-known disparities in genetic research ^101^, our analyses were limited to European ancestries, which do not allow generalization in all continental populations. We used MVP as the discovery cohort for EHR based CV diagnoses due to its relatively low sample overlap with PTSD GWAS, this may have filtered out some loci that could have been identified with other cohorts as discovery. Additionally, veteran participants may not be representative of genetic profile identified from other (civilian) populations due to underlying demographic differences. While our observational analysis in the AllofUs cohort confirmed the PTSD-CV comorbidities in civilian cohort, further studies will be needed to verify the same for PTSD-CV shared loci identified in our molecular analyses. The drug-gene targets and subsequent mapping to PTSD and CV conditions is explained as an example to learn about role of gene drugs in therapeutic and adverse effects of drugs. The drugs mentioned were identified from research studies and may not reflect actual prescription practices and need further validation before we can make hypotheses regarding their impact on patients’ implications. Pharmacovigilance data for gene-drug targets was extracted from a single source and may not be updated from research data or ongoing trials.

In conclusion, this study examines the shared biology of PTSD and CV health, leveraging large-scale molecular data and multi-modal information. Specifically, considering EHR diagnoses, cardiac imaging phenotypes, and cardiac health-related habits, we highlighted the local genetic correlations, proteomic and transcriptomic associations, and the potential pharmacological targets for molecular mechanisms underlying PTSD-CVD comorbidity. These finding converge on overlap in several domains across PTSD and CVD and within the context of the broader PTSD-CVD literature highlight important mechanisms relevant to potential pharmacological intervention warranting additional research.

## 5 Data Availability and Links

- UCSC Genome Browser : https://genome.ucsc.edu/cgi-bin/hgTables
- DLPFC Proteome weights: https://doi.org/10.7303/syn23627957
- Open Targets: https://genetics.opentargets.org/
- Pan-UKBB Summary Statistics: https://pan.ukbb.broadinstitute.org/
- MVP Summary Statistics: https://www.ncbi.nlm.nih.gov/projects/gap/cgi-bin/study.cgi?study_id=phs001672.v11.p1
- LAVA: https://github.com/josefin-werme/LAVA
- ARIC - plasma protein: http://nilanjanchatterjeelab.org/pwas/
- UKBB-PPP: https://metabolomips.org/ukbbpgwas/
- Phecode Map: https://phewascatalog.org/
- Meta-analyzed MVP-UKBB GWAS summary statistics of 14 traits: released at the time of publication.

## Data Availability

All data produced in the present work are contained in the manuscript. All data produced in the present study are available upon reasonable request to the authors.

## 6 Acknowledgements

This study was supported by grants from the National Institutes of Health (RF1 MH132337, R33 DA047527, and K99 AG078503), One Mind, the Alzheimer’s Association (Research Fellowship AARF-22-967171), Horizon 2020 (Marie Sklodowska-Curie Individual Fellowship 101028810), Yale Women’s Faculty Forum, and Suzhou Municipal Health Commission (LCZX202207). We also acknowledge the contribution of the participants and the investigators involved in the UK Biobank, the Million Veteran Program, the All of Us Research Program, and the Psychiatric Genomics Consortium. Major financial support for the PTSD-PGC was provided by the Cohen Veterans Bioscience, Stanley Center for Psychiatric Research at the Broad Institute, and the National Institute of Mental Health (NIMH; R01MH106595, R01MH124847, R01MH124851). The All of Us Research Program is supported by the National Institutes of Health, Office of the Director: Regional Medical Centers: 1 OT2 OD026549; 1 OT2 OD026554; 1 OT2 OD026557; 1 OT2 OD026556; 1 OT2 OD026550; 1 OT2 OD 026552; 1 OT2 OD026553; 1 OT2 OD026548; 1 OT2 OD026551; 1 OT2 OD026555; IAA #: AOD 16037; Federally Qualified Health Centers: HHSN 263201600085U; Data and Research Center: 5 U2C OD023196; Biobank: 1 U24 OD023121; The Participant Center: U24 OD023176; Participant Technology Systems Center: 1 U24 OD023163; Communications and Engagement: 3 OT2 OD023205; 3 OT2 OD023206; and Community Partners: 1 OT2 OD025277; 3 OT2 OD025315; 1 OT2 OD025337; 1 OT2 OD025276.

## 7 COMPETING INTERESTS

Dr. Polimanti reports a research grant from Alkermes outside the scope of this study. Drs. Polimanti and Gelernter are paid for their editorial work on the journal Complex Psychiatry. Dr. Gelernter is named as an inventor on PCT patent application no. 15/878,640 entitled “Genotype-guided dosing of opioid agonists,” filed January 24, 2018. Dr. Stein has in the past 3 years received consulting income from Actelion, Acadia Pharmaceuticals, Aptinyx, atai Life Sciences, Boehringer Ingelheim, Bionomics, BioXcel Therapeutics, Clexio, Delix Pharmaceuticals, EmpowerPharm, Engrail Therapeutics, GW Pharmaceuticals, Janssen, Jazz Pharmaceuticals, and Roche/Genentech; has stock options in Oxeia Biopharmaceuticals and EpiVario; and has been paid for editorial work on Depression and Anxiety (Editor-in-Chief), Biological Psychiatry (Deputy Editor), and UpToDate (Co-Editor-in-Chief for Psychiatry).Dr. O’Donnell an employee of Novartis Pharmaceuticals). Dr. Koller is the founder and CEO of EndoCare Therapeutics, but the company conducts research unrelated to the present study. The other authors declare no competing interests.

Supplementary Tables – See attached xlsx file.

---

**Supplementary Figure 1:**
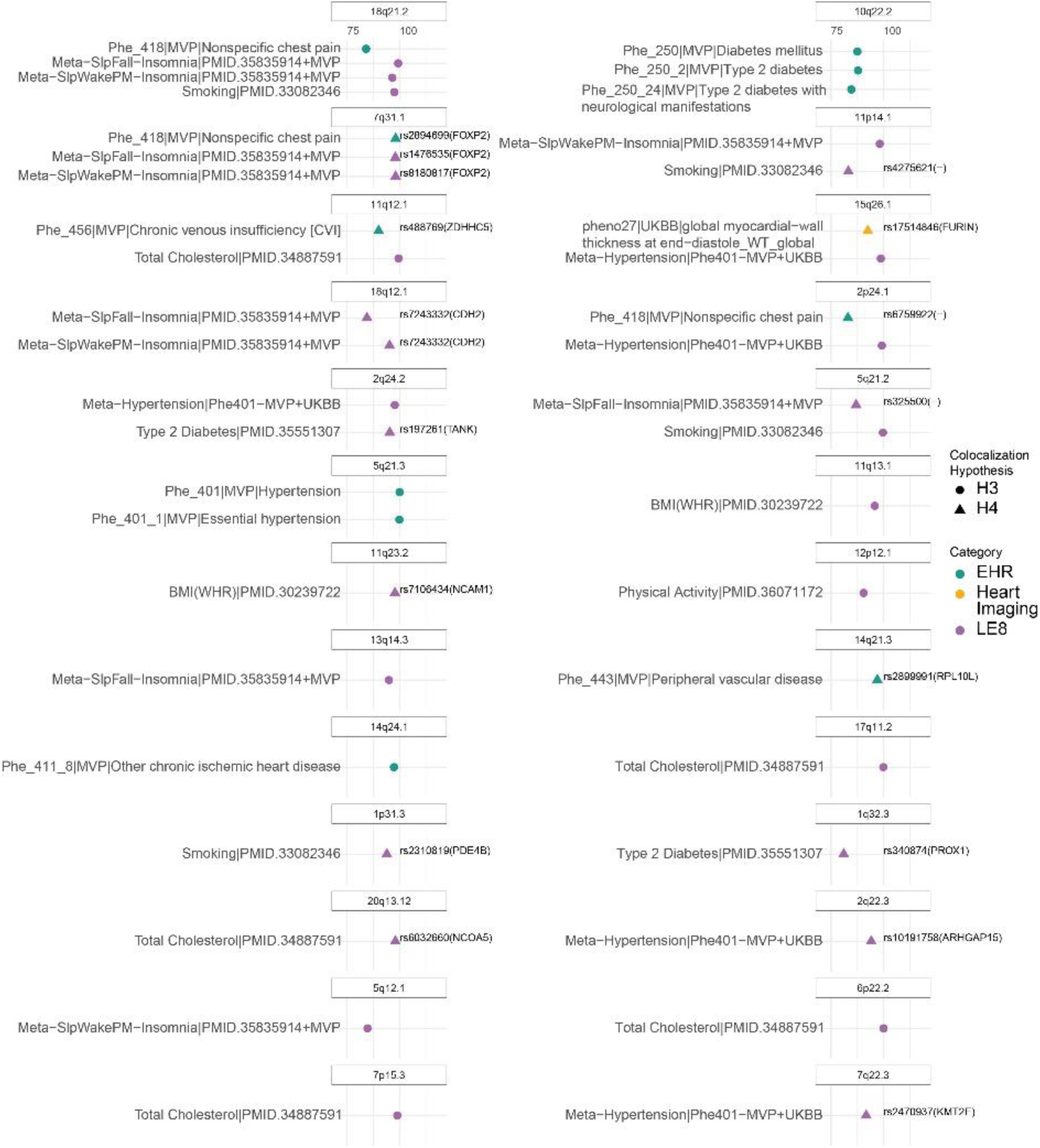
The x-axis shows colocalization probability(different LD-linked causal variants-H3 hypothesis; same causal variant – H4 hypothesis), and y-axis is marked with CV conditions. between PTSD and CV conditions based on colocalization. The shared causal variants based on H4 hypothesis are labelled and their nearest genes are in parenthesis. We highlight genes at each locus that had colocalization evidence with molecular profiles (gene/splicing/proteome expression) and CV conditions (Supplementary Table S7).

**Supplementary Figure 2:**
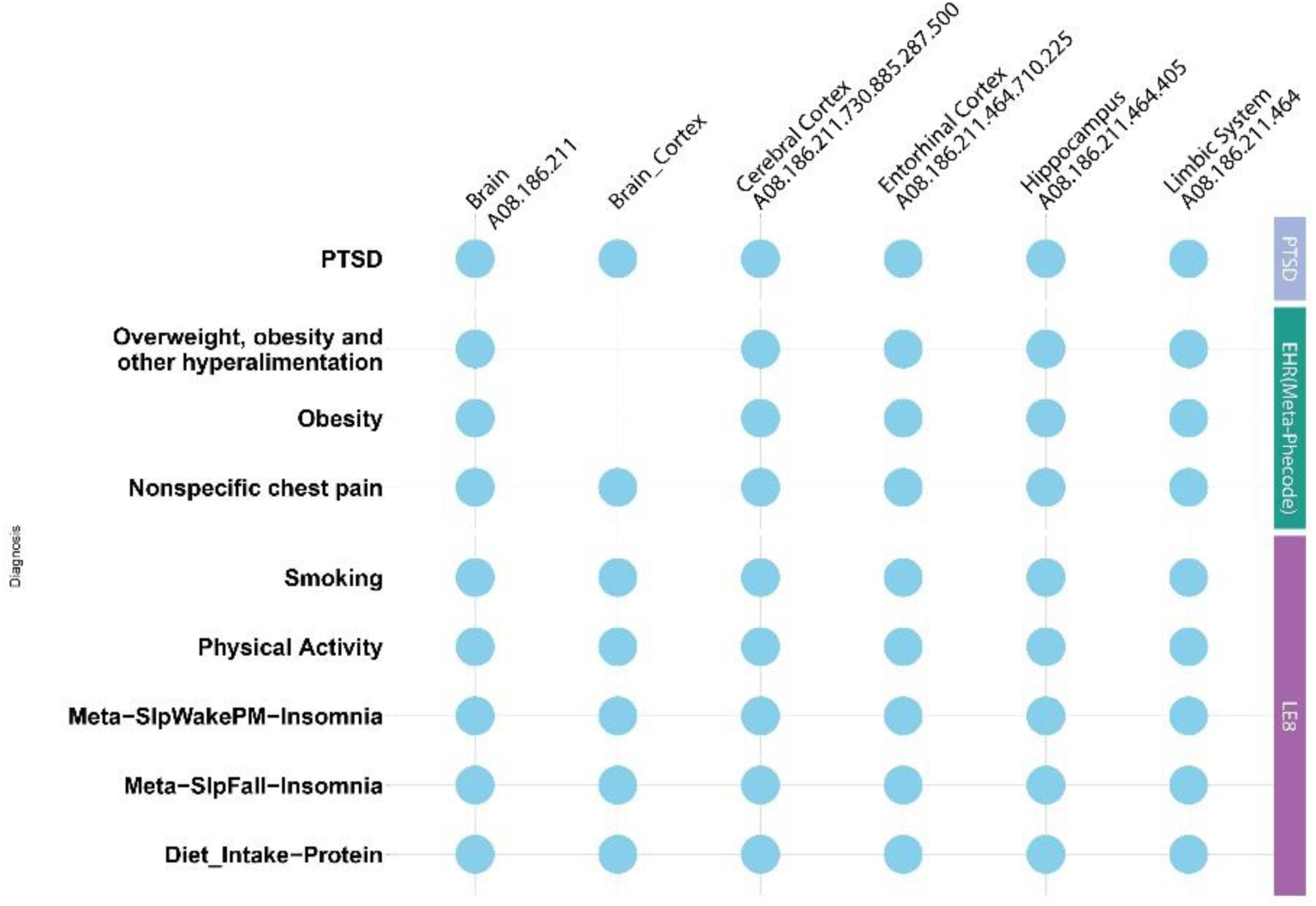
The matrix plot shows significant over-represented tissue based on genetically regulated gene expression profiles shared between PTSD and the CV conditions. The tissues are shown on the top x-axis, and traits are on the y-axis, grouped by category. LE8 – AHA’s Life’s Essential 8. Meta-analyzed phecodes from EHR of MVPandUKBB.

**Supplementary Figure 3:**
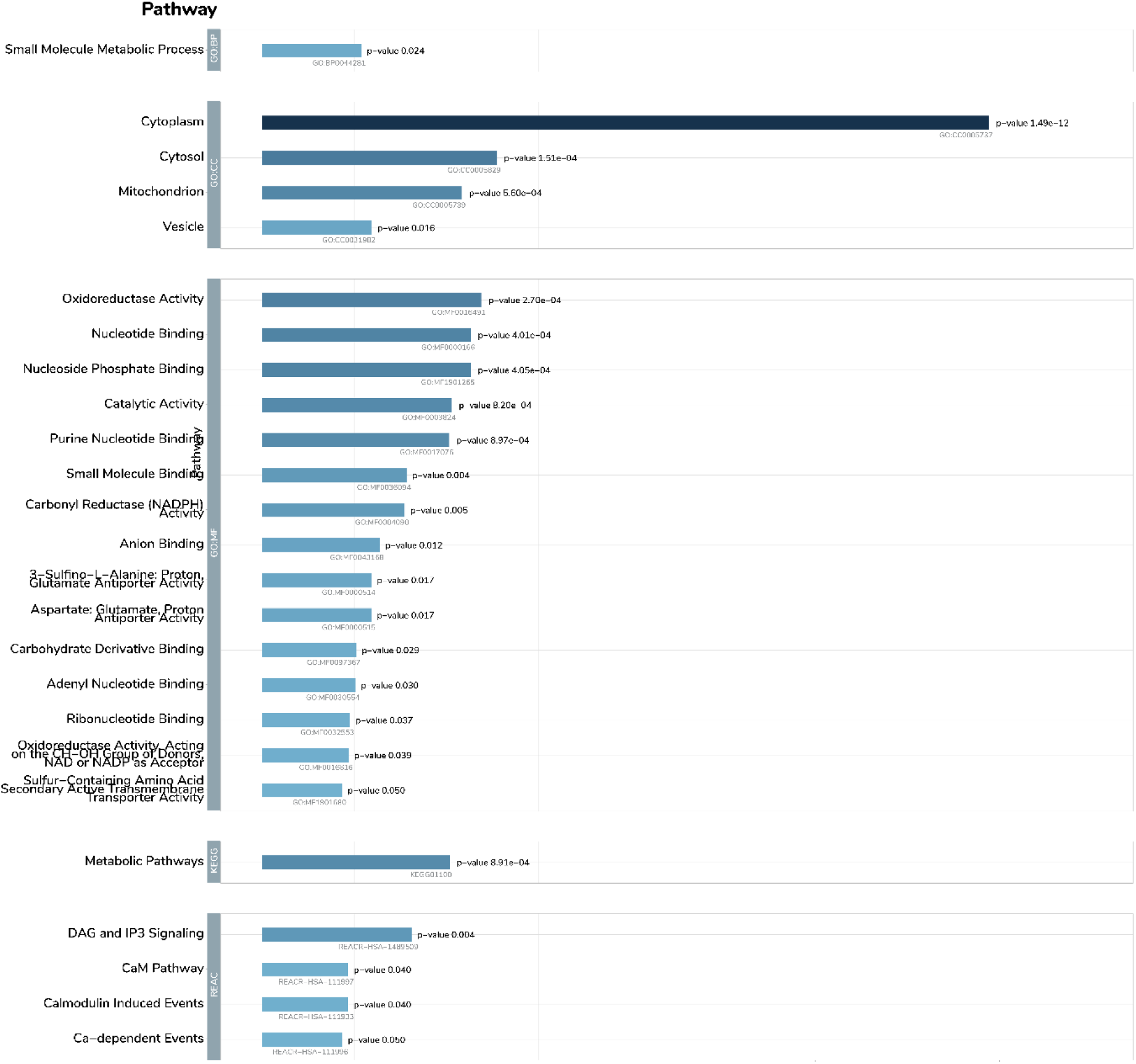
Bar plot showing pathway enrichment of genes overlapping between PTSD and CV conditions based on brain-based genetically regulated gene expression.

**Supplementary Figure 4:**
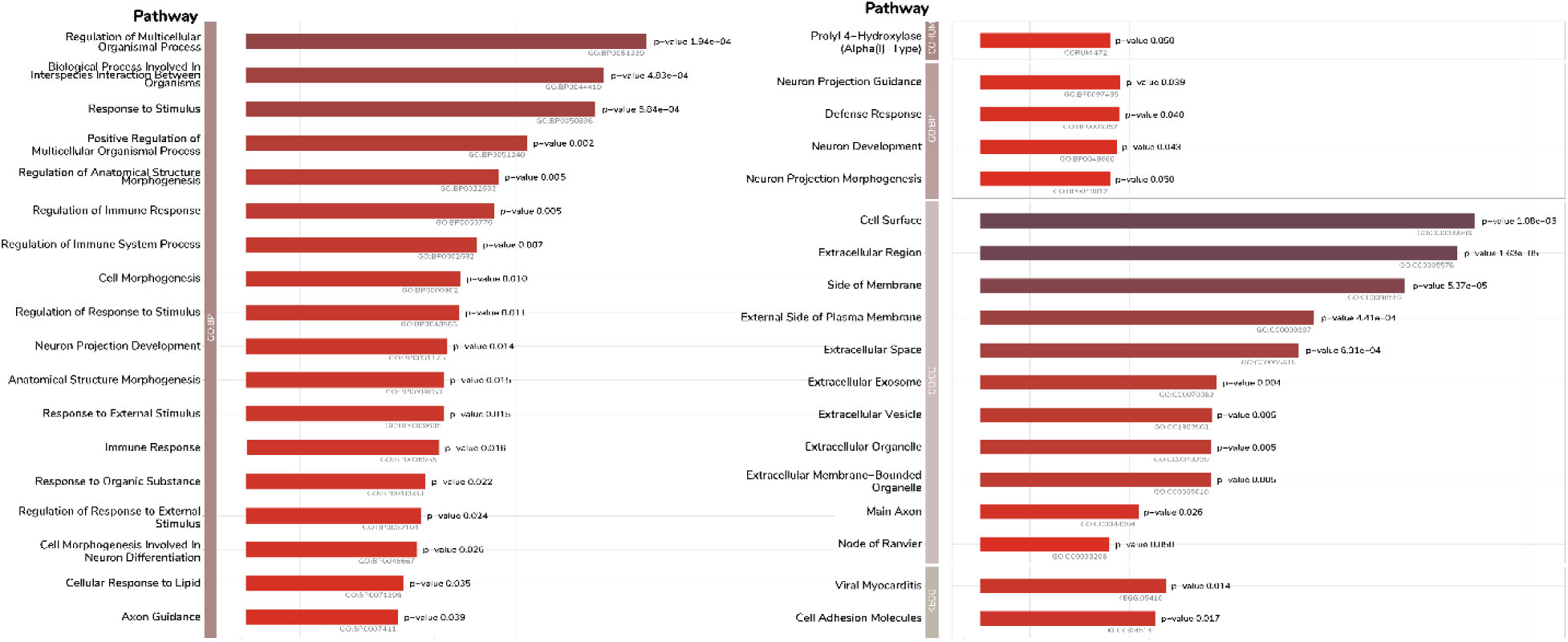
Bar plot showing pathway enrichment of genes overlapping between PTSD and CV conditions based on blood-based genetically regulated gene expression.

